# A signal processing tool for extracting features from arterial blood pressure and photoplethysmography waveforms

**DOI:** 10.1101/2024.03.14.24304307

**Authors:** R. Pal, A. Rudas, S. Kim, J.N. Chiang, M. Cannesson

## Abstract

Arterial blood pressure (ABP) and photoplethysmography (PPG) waveforms contain valuable clinical information and play a crucial role in cardiovascular health monitoring, medical research, and managing medical conditions. The features extracted from PPG waveforms have various clinical applications ranging from blood pressure monitoring to nociception monitoring, while features from ABP waveforms can be used to calculate cardiac output and predict hypertension or hypotension. In recent years, many machine learning models have been proposed to utilize both PPG and ABP waveform features for these healthcare applications. However, the lack of standardized tools for extracting features from these waveforms could potentially affect their clinical effectiveness. In this paper, we propose an automatic signal processing tool for extracting features from ABP and PPG waveforms. Additionally, we generated a PPG feature library from a large perioperative dataset comprising 17,327 patients using the proposed tool. This PPG feature library can be used to explore the potential of these extracted features to develop machine learning models for non-invasive blood pressure estimation.

## I. Introduction

Arterial blood pressure (ABP) serves as a fundamental hemodynamic parameter widely used to monitor and guide therapeutic interventions, especially in critically ill patients [1, 2]. The ABP waveform contains rich information about the cardiovascular system, including heart rate, systolic blood pressure, diastolic blood pressure, and mean arterial pressure [3]. On the other hand, Photoplethysmography (PPG), also known as the pulse oximetric wave, is a non-invasive method primarily employed in anesthetic monitoring for assessing blood oxygen levels (SaO2) [4]. The PPG waveform also carries rich information about cardiac activity and cardiovascular condition [5]. A variety of wearable devices based on PPG have been proposed to monitor heart rate, including smartphones and smartwatches [5].

In the context of analyzing ABP and PPG waveforms, a cardiac cycle is defined by five key points: systolic phase onset, systolic phase peak, dicrotic notch, diastolic phase peak, and diastolic phase endpoint (see Fig. 1). These key points serve as critical landmarks for extracting valuable features that have been used in various clinical applications. For example, as mentioned in [6], the duration from systolic phase onset to dicrotic notch, known as systolic phase duration, in the ABP waveform, can be used to monitor cardiac function. Additionally, as noted in [7], changes in dicrotic notch-based features, such as dicrotic notch amplitude and the systolic phase duration, often occur early in the vascular disease course and may help with early recognition. In PPG waveforms, the stiffness index, which is the ratio of the subject’s height to the duration between systolic phase peak and diastolic phase peak, has shown a relationship with the risk of coronary heart diseases, such as hypertension and diabetes [8, 9]. Moreover, the reflection index, which is the ratio of diastolic phase peak amplitude to systolic phase peak amplitude relative to systolic phase onset, serves as a valuable indicator for vascular assessment [8,10]. Additionally, the augmentation index, which is the ratio of the difference between systolic phase peak and diastolic phase peak amplitudes to the amplitude of systolic phase peak (with amplitudes relative to systolic phase onset), tends to increase in older individuals and those with cardiovascular disease [8,11].

**Fig. 1.**
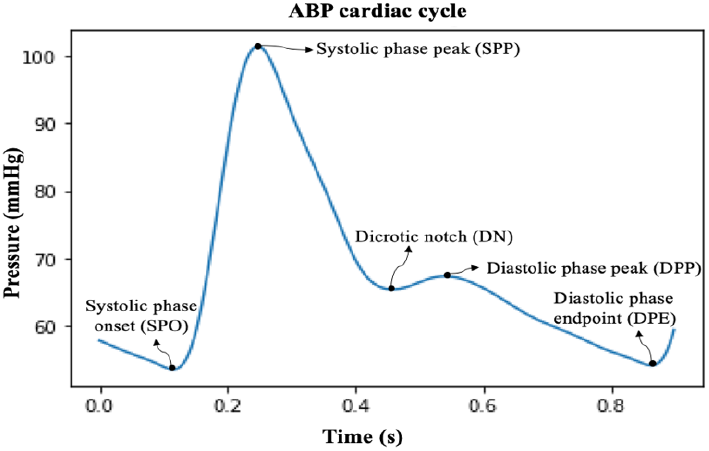
ABP cardiac cycle with all five key points: Systolic phase onset (SPO), Systolic phase peak (SPP), Dicrotic notch (DN), Diastolic phase peak (DPP), and Diastolic phase endpoint (DPE).

Many research groups have proposed machine learning models for non-invasive and continuous blood pressure measurement based on PPG waveform features as reported in [12,13]. Additionally, Hatib et al. [14] recently introduced a machine learning algorithm based on ABP features to predict hypotension.

These medical applications of PPG and ABP features clearly emphasize the importance of accurately detecting all five key points within a cardiac cycle in these waveforms, as they form the foundation for feature extraction and subsequent clinical applications. The use of derivatives (1^st^ and 2^nd^) of ABP and PPG waveforms is a common method for detecting key points within a cardiac cycle in these waveforms [15]. However, the sensitivity of signal derivatives to noise presents a significant challenge for accurate detection of these key points, potentially resulting in the extraction of inaccurate features.

Moreover, despite the growing significance of PPG and ABP features in medical applications, researchers currently lack a standard library of these features. This absence limits the research community’s collective ability to thoroughly explore their clinical utility in healthcare.

The main contributions of this paper are divided as follows:

- This study presents a signal processing tool based on the iterative envelope mean (IEM) method [16], that can detect all key points within cardiac cycle in ABP and PPG waveforms and can be used for extracting features from these waveforms.
- A PPG feature library (632 features/ cardiac cycle) along with simultaneous systolic blood pressure, diastolic blood pressure, and mean arterial pressure values extracted from corresponding cardiac cycles in the ABP waveform, was obtained from the large perioperative MLORD dataset comprising 17,327 patients [17], using the proposed tool.

## II. Methods

The process of detecting the temporal location of all five key points within a cardiac cycle in ABP/PPG signals and extracting features using these identified key points is referred to a signal processing tool for extracting features from ABP and PPG waveforms. The key points were detected using the iterative envelope mean method [16]. We recently introduced the iterative envelope mean fractal dimension filter for the separation of pulmonary crackles from normal breath sounds [16]. In this study, the concept of the iterative envelope mean (IEM) method is adapted for detecting the temporal location of all five key points within a cardiac cycle in ABP and PPG waveforms, serving as critical landmarks for feature extraction in these waveforms. The process is shown schematically in Fig. 2. The signal processing tool was developed using PYTHON programming language.

**Fig. 2.**
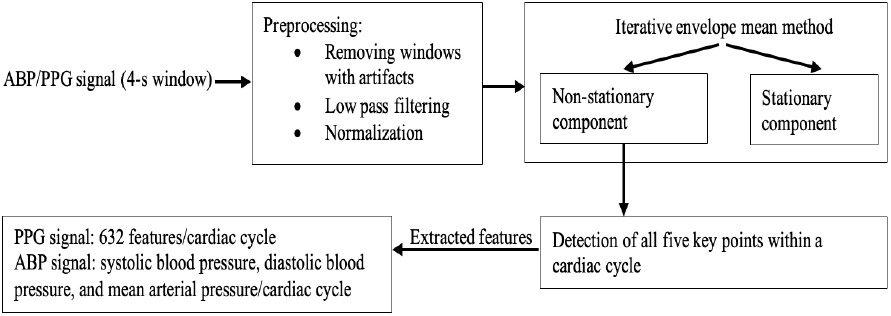
Block diagram of the signal processing tool using iterative envelope mean method for feature extraction.

### A. Pre-processing

The input consists of 4-s window containing ABP or PPG waveform. Windows that exhibit artifacts are excluded based on the following criteria: any window containing ‘0’ or negative values and having a number of peaks less than 3 or more than 10, exceeding 75^th^ percentile of the 4-s window amplitude. After artefacts removal a 4^th^-order Butterworth low-pass filter with a cutoff frequency of 16 Hz is applied to the 4-s input window to eliminate high-frequency noise. Additionally, the filtered signal is normalized using Eq. 1.

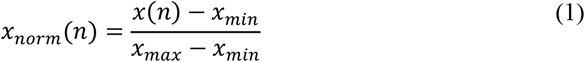

where *x*(*n*) represents the input signal, *n* is the sample index, *x*_*min*_ is the minimum value of the input signal, *x*_*max*_ is its maximum value, and *x*_*norm*_(*n*) denotes the normalized input signal.

### B. Iterative Envelope Mean Method

The IEM method decomposes the signal into its non-stationary and stationary components [16]. A 4-s window of the ABP input signal is shown in Fig. 3 (a). The IEM method procedure for a given input signal *x*(*n*) can be summarized as follows:

**Fig. 3.**
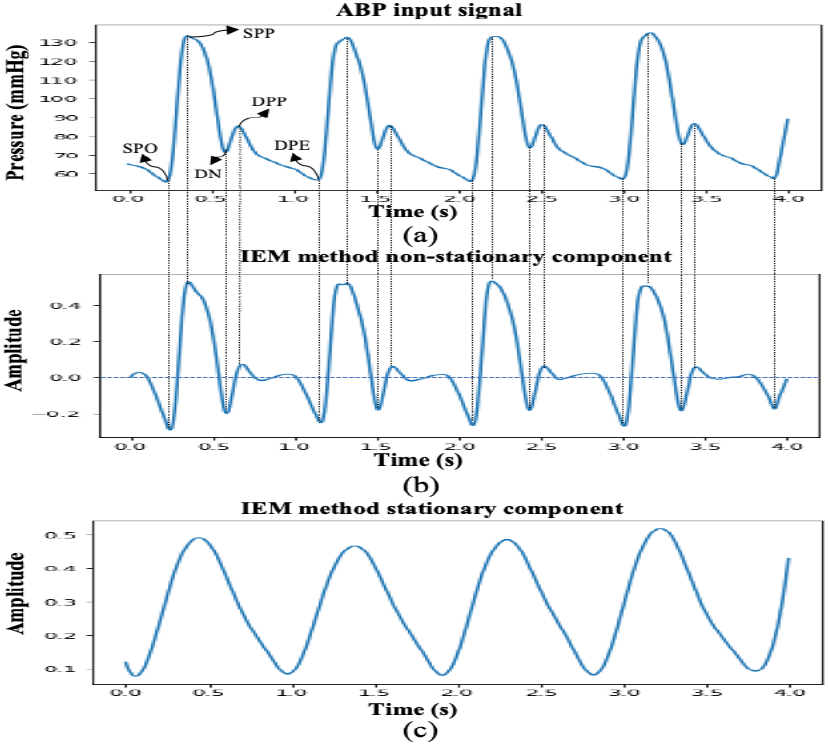
An example of the IEM method; (a) 4-s window of ABP input signal, (b) IEM method non-stationary component, and (c) IEM method stationary component. SPO: Systolic phase onset; SPP: Systolic phase peak; DN: Dicrotic notch; DPP: Diastolic phase peak; DEP: Diastolic end point.

Initially, the input signal is smoothed and its first and second derivatives are calculated using the Savizky-Golay (SG) family filter. The SG filter parameters used here are degree of fitting polynomial *p*_*f*_ =4, number of coefficients *n*_*c*_ =25 and order of derivation (*d*_*o*_) =0, 1 and 2 for smoothing the input signal, and for estimating the first and second derivatives of that smoothed signal, respectively. Note that as mentioned in [16] the number of coefficients *n*_)_, is approximately equal to the one to two times the half width of the shortest duration feature of interest in the signal. In the case of ABP/PPG signal the length of a cardiac cycle is approximately 0.8 s and the distance between two consecutive key points is 0.2 s. In our study, the sampling frequency is 256, therefore the half width is 25 samples and the value of *n*_*c*_ is equal to 25.

Now, the upper and lower envelopes are calculated using the coordinates of the smoothed input signal at the location of the first derivative local maxima and minima, respectively. The first derivative local extrema are calculated and classified as maxima and minima using the sign changes over the second derivative of the smoothed input signal. Then, an envelope mean value is determined by averaging the upper and lower envelopes of a smoothed input signal (Eq. 2).

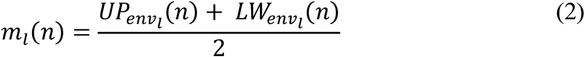

where *n* is the sample index in the input signal i.e. *n* =1, 2, …., *N* and *l* is the iteration number where *l*=1, 2,…, *L*.

The estimated envelope mean value is then subtracted from the original input signal and the resulting signal *R*_*l*_(*n*) is used as the input for subsequent iterations.

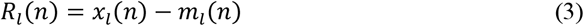

where *x*_*l*_(*n*) is the input signal at iteration *l*.

The iteration process ends when the stopping criterion is met (Eq. 4), at iteration *L*.

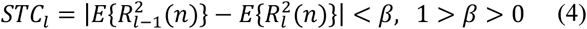

where E{.} denotes the expected value and has an initial value of *R*_*l*-*1*_ = 0. In this study, we have used accuracy level *β* = 0.*1*, and note that the IEM method employs the identical stopping criterion to that specified in [16,18].

After the last iteration (*L*) the IEM method provides an estimate of the non-stationary component (*NSTS*(*n*)) of the input signal (Eq. 5). Additionally, by summing up the envelope means from each iteration, it yields an estimate of the stationary component (*STS*(*n*)) of the signal (Eq. 6).

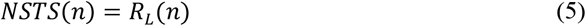

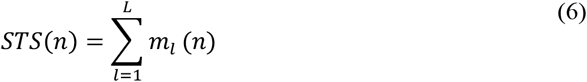

The non-stationary and stationary components after applying the IEM method are shown in Fig. 3(b) and (c), respectively. Although, the IEM method can reveal all key points within a cardiac cycle in its non-stationary component, several further steps are required for accurately locate the temporal location of these key points within a cardiac cycle in the non-stationary component. Challenges arise due to pressure reflections in the arterial system [19] and non-physiological oscillations in PPG and ABP waveforms, introducing multiple valleys within the cardiac cycle in NSTS that may lead to misinterpretation. To address this concern and precisely locate key points within a cardiac cycle, the following conditions are applied: (1) dicrotic notch and diastolic phase peak key points must be at least 0.1 s (25 samples) away from systolic phase peak and diastolic phase endpoint key points. (2) In the NSTS, the y-axis value for systolic phase peak point and diastolic phase peak point must be greater than zero and for systolic phase onset, dicrotic notch, and diastolic phase endpoint valleys, it must be less than zero.

### C. Data

A large perioperative dataset comprising 17, 327 patients (MLORD dataset) who underwent surgeries between 2019 and 2022 at the David Geffen School of Medicine at the University of California Los Angeles is used for the analysis [17]. The MLORD dataset includes both clinical data and waveform data. Clinical data were collected from Electronic Health Records (EHR), including Epic (Verona, WI, USA), and Surgical Information Systems (Alpharetta, GA, USA). The waveform data were collected in the operating room directly through the Bernoulli data collection system (Cardiopulmonary, New Haven, CT, USA) [17]. The waveform data spans more than 72,264 hours in time and is 7.6 TB in size, comprising various sampled digital physiological waveforms, such as PPG, ABP, and electrocardiogram. We refer readers to [17], for the detailed description of the MLORD dataset.

In the MLORD dataset, out of 17,327 patients, 4901 patients have ABP waveforms, 17170 patients have PPG waveforms, and 4893 patients have both ABP and PPG waveforms. In this study, we utilized the 4893 patients having both ABP and PPG waveforms. The sampling frequency of the ABP and PPG waveforms was 256 Hz. It is important to note that marking the temporal location of key points on all cardiac cycles as a reference is not feasible due to the dataset’s large size. Additionally, in some cases, either the dicrotic notch or diastolic phase peak is less pronounced, especially in PPG waveforms, making it difficult to establish their temporal location as a reference.

Therefore, to assess the performance of the proposed tool in terms of detecting key points within a cardiac cycle, 1000 4-s windows were randomly selected from both the ABP and PPG waveforms, where all five key points could be observed within a cardiac cycle. The ABP windows contained 3420 cardiac cycles, and the PPG windows consisted of 3440 cardiac cycles. An experienced researcher marked the key points within these cardiac cycles using the ‘find_peaks’ function from the scipy PYTHON package. To ensure accuracy, the marking was validated by an engineer and an anesthesiologist. They conducted a visual examination of marked ABP and PPG windows.

Moreover, to create a feature library we utilized 1,487,955 PPG cardiac cycles and an equal number of ABP cardiac cycles from 4893 patients with having both PPG and ABP waveforms in the MLORD dataset.

### D. Performance Evaluators

To evaluate the performance of the tool in terms of key points detection, three parameters: sensitivity (SE), positive predictive value (PPV), and F-score (*F*.) are used, as shown in Eq. (7), Eq. (8), and Eq. (9), respectively.

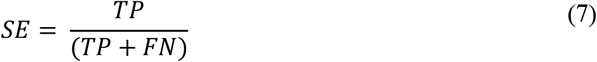

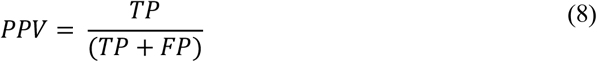

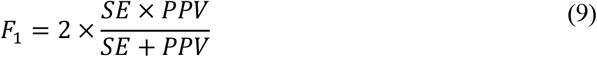

where TP stands for the number of true positives, FN for the number of false negatives, and FP for the number of false positives. Therefore, SE indicates the fraction of true (marked by experienced researcher) key points detected by the tool. PPV is the fraction of points assigned as key points by the tool which are true key points, and F_1_ is the harmonic mean of SE and PPV, commonly referred as a measure of the overall performance. Note that a threshold 8 ms (2 samples) was used to admit the proposed tool results as TP or reject them as FP or FN [20].

## III. Results and discussion

### A. Key points detection

The performance of the tool for detecting all five key points within a cardiac cycle in ABP and PPG waveforms is presented in Table 1. From Table 1, it can be observed that the proposed tool had, on average, a sensitivity (SE) of 100%, a positive predictive value (PPV) greater than 99 %, and an F_1_ score greater than 99 % for the detection of all key points within a cardiac cycle in both ABP and PPG waveforms. The tool utilizes a single signal (the non-stationary component of the IEM method) to identify the temporal location of all key points within a cardiac cycle, in contrast to the two different signals employed in the literature (1^st^ and 2^nd^ derivatives) [15]. Moreover, in terms of computational cost, the IEM method requires only *O* (*LN*) operations for number of iterations *L* and signal length of *N*. The graphical user interface we designed to help researchers and clinicians use our proposed tool is shown in Fig. 4. Furthermore, future research will focus on exploring the potential of the tool to detect the temporal location of key points when they are less distinct within a cardiac cycle, especially the dicrotic notch, as it tends to diminish with age [21]. Additionally, we will compare its performance against a multi-annotator gold standard with annotations verified by medical experts.

**Table 1.**
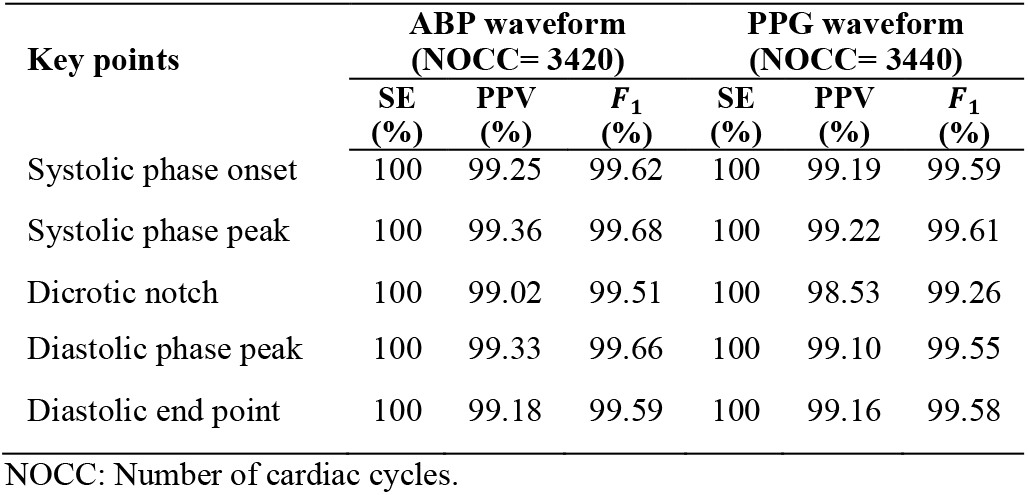
Overall performance of the signal processing tool for detecting all key points within CC in ABP and PPG waveforms.

**Fig. 4.**
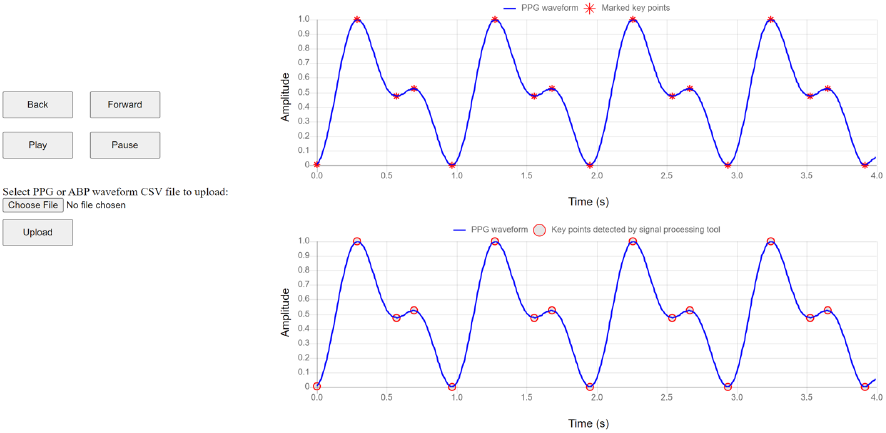
Graphical user interface (GUI) for key points detection within a cardiac cycle.

### B. Extracted features

After demonstrating excellent performance in detecting the temporal location of all five key points within a cardiac cycle on the randomly selected ABP and PPG cardiac cycles, the tool was applied to the complete set of PPG and ABP waveforms obtained from 4893 patients in the MLORD dataset. A total of 632 PPG features per cardiac cycle were extracted from the PPG waveforms, along with simultaneous systolic blood pressure, diastolic blood pressure, and mean arterial pressure values calculated from corresponding cardiac cycles in the ABP waveform. In total, 1,487,955 PPG cardiac cycles for extracting PPG features (632 features/PPG cardiac cycle) and an equal number of ABP cardiac cycles for calculating blood pressure values (systolic blood pressure, diastolic blood pressure, and mean arterial pressure/ABP cardiac cycle) were used to create the feature library. The extracted features from PPG cardiac cycles includes amplitude features (n_1_=30), amplitude ratio features (n_1_=210), duration features (n_1_=10), duration ratio features (n_1_=46), average features (n_1_=40), median features (n_1_=20), root mean square features (n_1_=20), area features (n_1_=40), area ratio features (n_1_=180), as well as systolic rise phase width and overall decay phase width features (n_1_=36), measured at different percentages of a cardiac cycle in the PPG waveform. For better understanding, extracted duration features are shown in Fig. 5(a) with their descriptions in Table 2. Moreover, a few extracted systolic rise phase width and overall decay phase width features are displayed in Fig. 5(b).

**Table 2.** Duration features (n_1_=10) description table.

| Feature index | Feature symbol | Description |
| --- | --- | --- |
| 1 | PPG_D_SPO_wrtSPP | Duration of PPG cardiac cycle systolic phase onset point with respect to systolic phase peak |
| 2 | PPG_D_SPO_wrtDN | Duration of PPG cardiac cycle systolic phase onset point with respect to dicrotic notch |
| 3 | PPG_D_SPO_wrtDPP | Duration of PPG cardiac cycle systolic phase onset point with respect to diastolic phase peak |
| 4 | PPG_D_SPO_wrtDPE | Duration of PPG cardiac cycle systolic phase onset point with respect to diastolic phase end point |
| 5 | PPG_D_SPP_wrtDN | Duration of PPG cardiac cycle systolic phase peak point with respect to dicrotic notch |
| 6 | PPG_D_SPP_wrtDPP | Duration of PPG cardiac cycle systolic phase peak point with respect to diastolic phase peak |
| 7 | PPG_D_SPP_wrtDPE | Duration of PPG cardiac cycle systolic phase peak point with respect to diastolic phase end point |
| 8 | PPG_D_DN_wrtDPP | Duration of PPG cardiac cycle dicrotic notch point with respect to diastolic phase peak |
| 9 | PPG_D_DN_wrtDPE | Duration of PPG cardiac cycle dicrotic notch point with respect to diastolic phase end point |
| 10 | PPG_D_DPP_wrtDPE | Duration of PPG cardiac cycle diastolic phase peak point with respect to diastolic phase end point |

**Fig. 5.**
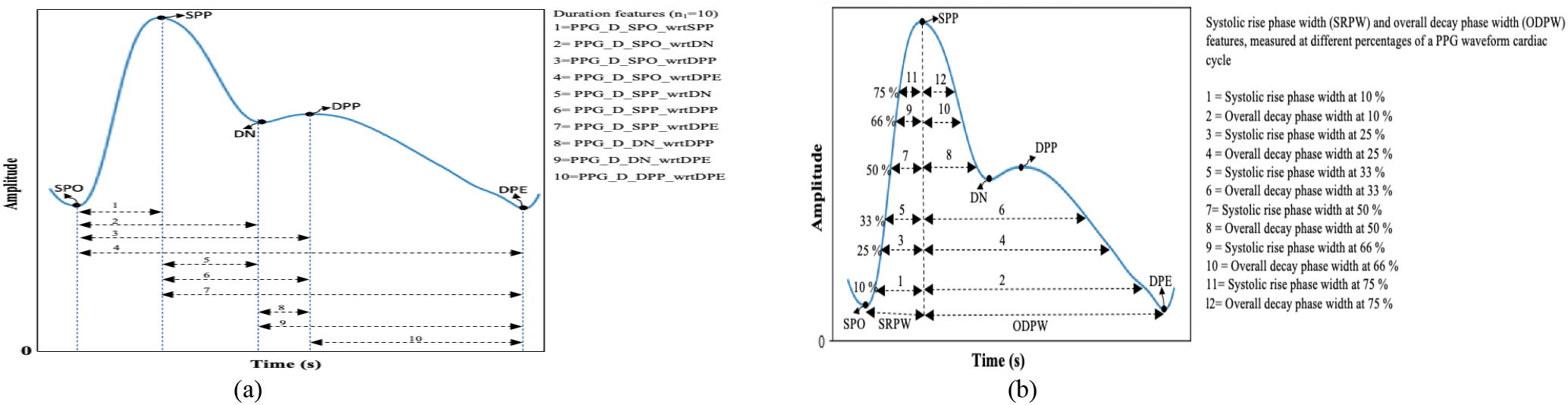
PPG cardiac cycle features; (a) Duration features, (b) Systolic rise phase width and overall decay phase width features.

## IV. Conclusion

We conclude that our proposed signal processing tool can be utilized for extracting features from ABP and PPG waveforms. The potential of these features can then be explored in feature-based machine learning models for non-invasive blood pressure estimation and the prediction of hypotension or hypertension. Additionally, the developed feature library, which includes extracted features from PPG waveforms along with the blood pressure values (systolic blood pressure, diastolic blood pressure, and mean arterial pressure), provides a valuable resource for researchers worldwide to explore the feasibility of PPG features in developing machine learning models for non-invasive blood pressure estimation. Future research will focus on validating the noise robustness performance of the proposed tool in terms of key points detection and expanding the feature library by integrating frequency domain features derived from PPG waveforms.

## Data Availability

The interested parties may contact the first author at or the corresponding author at to request access to the MLORD dataset, developed feature library with the detailed description (supplementary material file) of extracted features, and proposed featurization tool.

## Declaration of competing interest

Dr. Cannesson is a consultant for Edwards Lifesciences and Masimo Corp, and has funded research from Edwards Lifesciences and Masimo Corp. He is also the founder of Sironis and Perceptive Medical and he owns patents and receives royalties for closed loop hemodynamic management technologies that have been licensed to Edwards Lifesciences.

## Acknowledgment

This work was supported by the National Institutes of Health (NIH): R01EB029751 and 1R01HL144692.

## Notes

Research supported by the National Institutes of Health (NIH): R01EB029751 and 1R01HL144692

### Funding Statement

Research supported by the National Institutes of Health (NIH):R01EB029751 and R01HL144692

### Author Declarations

Institutional Review Board of University of California, Los Angeles gave ethical approval for this work.

## References

[1] M. A. Navakatikyan, C. J. Barrett, G. A. Head, J. H. Ricketts, and S. C. Malpas, “A real-time algorithm for the quantification of blood pressure waveforms,” IEEE Tran. on Biomedical Engineering, vol. 49, no. 7, pp. 662–670, July 2002.

[2] B. H. McGhee and E. J. Bridges, “Monitoring arterial blood pressure: What you may not know,” Crit. Care Nurse, vol. 22, no. 2, pp. 60–79, April 2002.

[3] W. Zong, T. Held, G. B. Moody, and R. G. Mark, “An open-source algorithm to detect onset of arterial blood pressure pulses,” in Proc. Comput. Cardiol., 2003, pp. 259–262.

[4] I. Korhonen and A. Yli-Hankala, “Photoplethysmography and nociception,” Acta Anaesthesiologica Scand., vol. 53, no. 8, pp. 975–985, 2009.

[5] T. Pereiraet, N. Tran, K. Gadhoumi et al., “Photoplethysmography based atrial fibrillation detection: a review,” Npj Digit. Med., vol. 3, no. 1, pp. 1–12, Jan. 2020.

[6] S. Hoeksel, J. Jansen, J. Blom, and J. J. Schreuder, “Detection of dicrotic notch in arterial pressure signals,” Journal of clinical monitoring, vol. 13, no. 5, pp. 309–316, 1997.

[7] A. Abushouk, T. Kansara, O. Abdelfattah et al., “The Dicrotic Notch: Mechanisms, Characteristics, and Clinical Correlations,” Curr Cardiol Rep, vol. 25, pp. 807–816, 2023.

[8] M. Z. Suboh, R. Jaafar, N. A. Nayan, N. H. Harun, and M. S. F. Mohamad, “Analysis on four derivative waveforms of photoplethysmogram (PPG) for fiducial point detection,” Frontiers in Public Health, vol. 10:920946, June 2022.

[9] V. V. S. Bonarjee, “Arterial stiffness: A prognostic marker in coronary heart disease. Available methods and clinical application,” Front. Cardiovasc. Med., vol. 5, Art. no. 64, 2018.

[10] J. M. Ahn, “New aging index using signal features of both photoplethysmograms and acceleration plethysmograms,” Healthcare informatics research, vol. 23, no. 1, pp. 53–59, 2017.

[11] U. Rubins, A. Grabovskis, J. Grube, and I. Kukulis, “Photoplethysmography analysis of artery properties in patients with cardiovascular diseases,” in Proc. 14th Nordic-Baltic Conf. Biomed. Eng. Med. Phys., 2008, pp. 319–322.

[12] C. El-Hajj and P. A. Kyriacou, “A review of machine learning techniques in photoplethysmography for the non-invasive cuff-less measurement of blood pressure,” Biomed. Signal Process. Control, vol. 58, Art. no. 101870, Apr. 2020.

[13] B. Tarifi, A. Fainman, A. Pantanowitz, and D. M. Rubin, “A Machine Learning Approach to the Non-Invasive Estimation of Continuous Blood Pressure Using Photoplethysmography,” Appl. Sci.,vol. 13,. no. 6:3955, March 2023.

[14] F. Hatib, Z. Jian, S. Buddi, C. Lee, J. Settels, K. Sibert, J. Rinehart, and M. Cannesson, “Machine-learning algorithm to predict hypotension based on high-fidelity arterial pressure waveform analysis,” Anesthesiology, vol. 129, no. 4, pp. 663–674, Oct. 2018.

[15] L. Peter, J. Kracik, M. Cerny, N. Noury, and S. Polzer. “Mathematical Model Based on the Shape of Pulse Waves Measured at a Single Spot for the Non-Invasive Prediction of Blood Pressure,” Processes, vol. 8, no. 4: 442, 2020.

[16] R. Pal, A. Barney, “Iterative envelope mean fractal dimension filter for the separation of crackles from normal breath sounds,” Biomedical Signal Processing and Control, vol. 66: 102454, 2021.

[17] S. Kim, S. Kwon, A. Rudas, R. Pal, M.K. Markey, A.C. Bovik, M. Cannesson, “Machine Learining of Physiologic Waveforms and Electronic Health Record Data: A Large Perioperative Data Set of High-Fidelity Physiologic Waveforms,” Crit Care Clin., 2023.

[18] L. J. Hadjileontiadis and S. M. Panas, “Separation of discontinuous adventitious sounds from vesicular sounds using a wavelet-based filter,” IEEE Trans. Biomed. Eng., vol. 44, no.12, pp. 1269–1281, Dec. 1997.

[19] M. Nirmalan, P.M. Dark, “Broader applications of arterial pressure wave form analysis,” Continuing Education in Anaesthesia, Critical Care & Pain, vol. 14, no. 6, pp. 285–290, 2014.

[20] N. Li, M. C. Dong, and M. I. Vai, “On an automatic delineator for arterial blood pressure waveforms, Biomed. Signal Process. Control, vol. 5, no. 1, pp. 76–81, Jan. 2010.

[21] J. Balmer, R. Smith, C.G. Pretty, T. Desaive, G.M. Shaw, J.G. Chase, “Accurate end systole detection in dicrotic notch-less arterial pressure waveforms,”, J Clin Monit Comput, p.p 1–10, 2020.

